# Network connectivity and local transcriptomic vulnerability underpin cortical atrophy progression in Parkinson’s disease

**DOI:** 10.1101/2023.04.20.23288538

**Authors:** Andrew Vo, Christina Tremblay, Shady Rahayel, Golia Shafiei, Justine Y. Hansen, Yvonne Yau, Bratislav Misic, Alain Dagher

## Abstract

Parkinson’s disease pathology is hypothesized to spread through the brain via axonal connections between regions and further modulated by local vulnerabilities within those regions. The resulting changes to brain morphology have previously been demonstrated in both prodromal and *de novo* Parkinson’s disease patients. However, it remains unclear whether the pattern of atrophy progression in Parkinson’s disease over time is similarly explained by network-based spreading and selective vulnerability. We address this gap by mapping the trajectory of cortical atrophy rates in a large, multicentre cohort of Parkinson’s disease patients and related this atrophy progression pattern to network architecture and gene expression profiles. Across 4-year follow-up visits, increased atrophy rates were observed in posterior, temporal, and superior frontal cortices. We demonstrated that this progression pattern was shaped by network connectivity. Regional atrophy rates were strongly related to atrophy rates across structurally and functionally connected regions. We also found that atrophy progression was associated with specific gene expression profiles. The genes most related to atrophy rates were those enriched for mitochondrial and metabolic function. Taken together, our findings demonstrate that both global and local brain features influence vulnerability to neurodegeneration in Parkinson’s disease.

## INTRODUCTION

Structural brain changes are observed in Parkinson’s disease (PD; [60, 92, 111]). The spatial topography of disease-related atrophy is not uniform across the cortex, however. Some regions appear to be more vulnerable to disease pathology than others, but the brain features that may determine this atrophic pattern and its progression are not entirely clear. One possibility is that PD occurs through a network spreading process. Early proposals by Braak et al., [16, 17] described a spatiotemporal distribution of Lewy pathology in post-mortem samples that followed a stereotyped caudal-to-rostral gradient. Pathology appeared first in the dorsal motor nucleus of the vagus in the lower brainstem, then progressed to the midbrain coinciding with the onset of motor symptoms that typify PD, before reaching subcortical and cortical areas. This pattern of disease spread appears to reflect brain network organization, suggesting that PD might target large-scale intrinsic networks [85, 105].

Recent evidence involving alpha-synuclein fibrils injected in mice further supports the theory that PD pathology propagates via neuronal connections [56, 65, 69]. MRI studies in PD patients are also consistent with a network spreading model. Brain atrophy in PD measured with deformation-based morphometry was found to correlate with connectivity to the substantia nigra, a presumed disease epicenter [111]. Cortical thinning in PD was associated with structural and functional connectivity to a subcortical “disease reservoir” [107]. The observed pathology distribution in PD does not always fit the pattern originally described by Braak’s staging model [15], however, which suggests that factors other than or in addition to network connectivity may shape the disease pattern.

One such factor might be the selective vulnerability of certain regions to PD pathology [35, 95], which can be conferred by local features such as cellular composition [106], neuroreceptor profiles [38], and gene expression [6]. For example, variations in regional alpha-synuclein protein expression have been shown to produce differences in vulnerability to neurodegeneration [57]. Agent-based simulations of pathology spread in PD also showed that incorporation of local differences in gene expression related to alpha-synuclein synthesis and degradation improved the *in silico* recreation of brain atrophy, suggesting a crucial involvement of regional gene expression in shaping disease propagation [77, 79, 113]. Indeed, the underlying factors that shape the atrophy pattern PD are complex and multifactorial, requiring further investigation.

In the present study, we aimed to characterize the network connectivity and local gene expression features underpinning the pattern of cortical atrophy progression in PD. We first charted the trajectory of cortical atrophy using the Parkinson Progression Marker Initiative (PPMI; [61]), a large, multi-centre cohort of *de novo* PD patients followed longitudinally up to 4 years after diagnosis. We then explored whether the cortical atrophy progression pattern mapped onto specific functional networks, cytoarchitectonic classes, cellular distributions, and neurotransmitter systems. Next, we applied a network spreading model [88, 89] to examine whether network connectivity constrained the atrophy progression pattern in PD.

Finally, we investigated the gene expression profiles associated with cortical atrophy progression in PD and its transcriptional relevance in terms of biological processes.

## MATERIALS AND METHODS

Clinical and imaging data used in this study are part of the PPMI database and can be accessed at http://www.ppmi-info.org/data. All other datasets and analysis tools along with their sources are cited in the *Materials and Methods* section.

### Participants

Demographic, clinical, and MRI data were downloaded from the PPMI (http://www.ppmi-info.org; [61]). All participating sites received approval from their local research ethics committee. Informed consent was obtained from all participants in accordance with the Declaration of Helsinki [8]. PD patients met the inclusion criteria outlined by the PPMI. Healthy controls were free of neurological disease. For participants with available 3T MRI data, only PD patients with a minimum of two visits and healthy controls with a baseline visit (due to limited longitudinal imaging in this group) were included in the present study.

### Cortical thickness analysis

T1-weighted MRI scans were acquired following standardized procedures and acquisition parameters (http://www.ppmi-info.org/study-design/research-documents-and-sops/). We used the CIVET 2.1.1 pipeline (http://www.bic.mni.mcgill.ca/ServicesSoftware/CIVET) to generate cortical thickness maps for PD patients across follow-up visits and healthy controls at baseline [3]. Briefly, T1-weighted MRIs were linearly transformed to the MNI-ICBM152 volumetric template, corrected for signal intensity non-uniformity, and segmented into grey matter, white matter, CSF, and background [27**?**, 28]. The grey and white matter surface boundary was fitted across 40,692 vertices in each hemisphere (inner cortical surface) and expanded to fit the grey matter and CSF boundary (outer cortical surface) using a fully automated method (Constrained Laplacian Anatomic Segmentation using Proximity algorithm; [47]). Cortical thickness at each of 81,920 vertices was measured as the linked distance between corresponding inner and outer surface vertices in millimetres. Quality control of the resulting outputs was carried out by two independent reviewers and only scans with consensus of the two reviewers were retained for further analysis. 146 of the 624 PD and 57 of the 194 HC scans failed quality control. Reasons for exclusion involved: motion artifacts, low signal-to-noise ratio, artifacts due to hyperintensities from blood vessels, surface-surface intersections, and poor grey and white matter segmentation.

To control for age- and sex-related effects on cortical thickness, we generated W-score maps for each PD patient at each follow-up visit. W-scoring is comparable to *z*-scoring but allows for the combined adjustment of many covariates [44]. Vertex-wise linear regressions were first performed in healthy controls between age, sex, and cortical thickness. W-score maps were then computed in PD patients using the following formula:

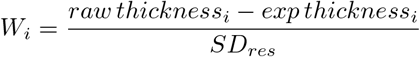

where *W*_*i*_ is the W-score at vertex *i, raw thickness*_*i*_ is the raw cortical thickness in PD measured at vertex *i, exp thickness*_*i*_ is the expected cortical thickness in healthy controls at vertex *i* given the patient’s age and sex (*thickness ∼ AGE* + *SEX*), and *SD*_*res*_ is the standard deviation of the residuals in healthy controls. Note that only data from healthy controls at baseline were used as the reference because only a limited number had longitudinal MRI data available. For interpretability, W-scores were inverted such that more positive scores indicated greater atrophy whereas more negative scores denoted lower atrophy. These W-scored maps were then used to model cortical atrophy rates with linear mixed effects.

Cortical atrophy maps were parcellated into equally sized regions (or parcels) using a multi-scale edition of the Desikan-Killiany atlas [24] referred to here as the “Cammoun atlas” [19]. Available parcellation resolutions spanned 68, 114, 219, 448, and 1000 cortical regions. Parcel-wise values were calculated as the mean value of all vertices assigned to a given parcel according to the Cammoun atlas. We modeled cortical thickness at different parcellation resolutions to ensure results were replicable and robust to the choice of spatial scale.

### Cortical atrophy progression analysis

W-scored vertex-wise cortical thickness values were averaged for the whole-brain as well as within parcels of the Cammoun atlas [19]. Linear mixed effects models were used to examine the change in cortical thickness across follow-up visits:

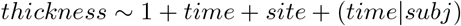

Time since baseline visit (years) and study site were used as fixed effects of interest [11]. Subject-wise slope and intercept were modeled as random effects. Separate models were fitted for each parcel and results were corrected for multiple comparisons using false discovery rate (FDR)-correction with *p* < 0.05 indicating statistical significance. The resulting t-statistic map corresponding to the effect of time on cortical thickness in PD was used as a measure of atrophy progression in all subsequent analyses.

### Clinical variable progression analysis

Analogous to our cortical atrophy progression analysis, we performed linear mixed effects modeling on longitudinal clinical data in PD patients. The full list of clinical variables is reported in Table 1. Imputation of missing scores (7.6%) was performed through replacement with the group mean at each time point. Separate models were fitted for each clinical outcome:

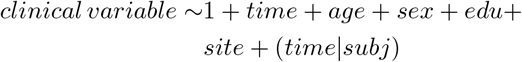

with time since baseline visit (years), adjusted for age, sex, education, and study site as fixed effects. Subject-specific slope and intercept were modeled as random effects. Results were FDR-corrected for multiple comparisons using a threshold of *p* < 0.05.

**TABLE 1:**
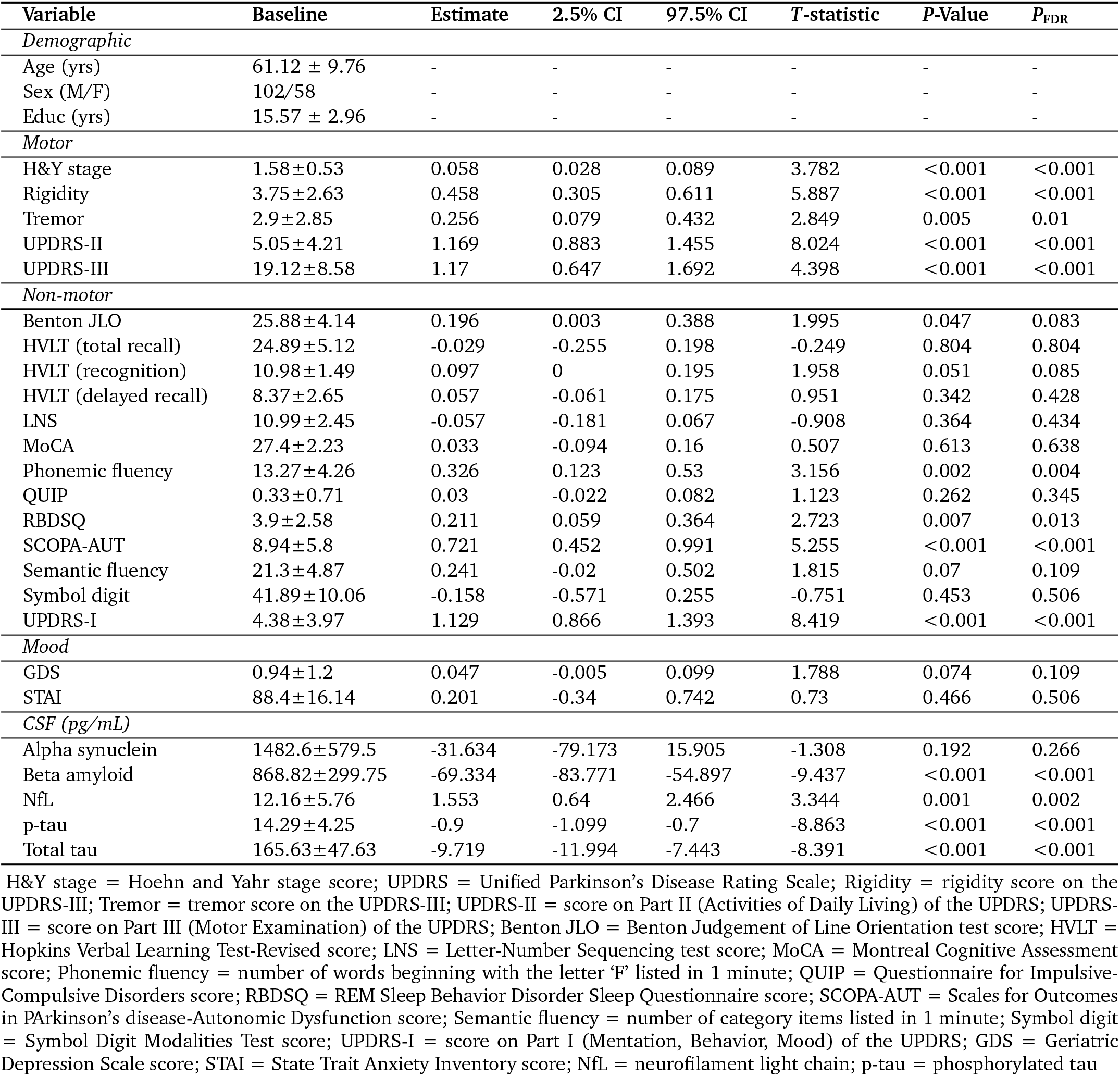
Demographic and clinical variable progression. Parkinson’s disease patient (*N* = 160) baseline demographics and ‘time’ effect parameter estimates for linear mixed effect models fit to longitudinal clinical data.

### Atrophy-clinical progression relationship

Behavioural partial least squares (PLS) analysis is a multivariate approach that identifies latent variables that explain the maximum covariance between two datasets [50, 66]. Here, we related a regional atrophy progression matrix *X*_*n×p*_ (160 patients × 68 regions) with a clinical progression matrix *Y*_*n*_ *×* _*q*_ (160 patients × 25 clinical outcomes). The matrices were first *z*-scored columnwise and used to generate an atrophy-clinical covariance matrix *R*_*q*_*×*_*p*_ = *Y* ^*0*^*X*. This covariance matrix was then subjected to singular value decomposition (SVD; [25]) such that *R* = *USV*′ where *U*_*p×l*_ and *V*_*q×l*_ are orthogonal matrices of left and right singular values and *S*_*l l*_ is a diagonal matrix of singular values. Each latent variable *i* was comprised of the *i*th columns of *U* and *V* and the covariance between these singular vectors was represented by the *i*th element of *S*. Left singular vectors of *U* represented the degree to which each brain region contributed to a latent variable (i.e., brain weight) whereas right singular vectors of *V* represented the degree to which clinical outcomes contributed to the same latent variable (i.e., clinical weight). Positively weighted brain regions covaried with positively weighted clinical outcomes and negatively weighted brain regions covaried with negatively weighted clinical outcomes. Pearson correlation tested the relationship between regional brain weights and atrophy rates, with a positive correlation indicating more positively weighted brain regions were associated with greater cortical atrophy progression. The effect size (*η*) associated with a given latent variable was estimated as the ratio of the squared singular value (*σ*) to the sum of all squared singular values. SVD generates the same number of latent variables as the rank of the covariance matrix *R* (i.e., 25).

Permutation tests were used to evaluate the statistical significance of each latent variable; that is, whether the variance explained was significantly greater than what would be expected by a null. Rows of matrix *X* were randomly reordered to generate a covariance matrix *R* from the permuted matrix *X* and original matrix *Y* before applying SVD. This procedure was repeated 5,000 times to generate a null distribution of singular values to which the original singular value was compared. Bootstrap resampling was then used to estimate the contribution and reliability of individual variables (i.e., brain regions and clinical outcomes) to each latent variable. The rows of both matrices *X* and *Y* were randomly selected with replacement to generate a resampled covariation matrix *R* that was subjected to SVD. This procedure was repeated 5,000 times to generate a sampling distribution for each weight in the singular vectors *U* and *V*. For each singular vector, a bootstrap ratio was calculated as the ratio of each singular vector to its bootstrap-estimated standard error. Thus, individual variables with larger bootstrap ratios had larger weights (i.e., contributed greatly to the multivariate pattern) and smaller standard errors (i.e., were more reliable).

### Null models

“Spin tests” [4, 64], or spatial autocorrelation-preserving null model testing, were used to assess the statistical significance whenever examining the correspondence between topologies of two brain maps. Given that neighbouring brain regions are not statistically independent and instead demonstrate a high degree of autocorrelation, null models that preserve this spatial autocorrelation of brain maps allow for more accurate inferences of the underlying data [64]. Null models were generated using the netneurotools toolbox [64]. Briefly, each parcel of the Cammoun atlas registered on the FreeSurfer fsaverage surface space was assigned the spatial coordinate of the vertex on an fsaverage spherical projection that was closest to its center of mass. The spatial coordinates were then randomly rotated for one hemisphere and mirrored to the other hemisphere. Lastly, each original parcel was reassigned the value of the closest rotated parcel. This procedure was repeated 10,000 times to generate a spatial autocorrelation-preserving null distribution against which empirical observations could be benchmarked [101].

### Spatial overlap between atrophy rates and brain features

We tested if the longitudinal atrophy pattern was associated with specific brain features such as intrinsic networks [109], tissue cytoarchitectonic classes [84, 104], cell type distributions [86, 90], and neurotransmitter receptor and transporter densities [38, 63]. The Yeo intrinsic networks [109] classify brain regions as one of seven resting state networks (visual, sensorimotor, dorsal attention, ventral attention, limbic, frontoparietal, and default mode) and the von Economo cytoarchitectonic classes [84] label brain regions as one of seven laminar cortical types (primary motor, association 1/2, primary/secondary sensory, primary sensory, limbic, and insular). We calculated the mean atrophy progression for each intrinsic network and cytoarchitectonic class and compared these empirical means to null mean distributions generated from spatial autocorrelation-preserving null models to determine their statistical significance.

For cell type distributions, we correlated the longitudinal atrophy pattern with the gene expression maps of seven different cell classes including: astrocytes, endothelial cells, excitatory and inhibitory neurons, microglia, oligodendrocytes, and oligodendrocyte precursors. Each cell class was first associated with a gene list derived from single-cell RNA sequencing studies of postmortem human cortical samples [86]. The spatial expression patterns of each gene list were generated and parcellated to the Cammoun atlas using post-mortem brain data from the Allen Human Brain Atlas (AHBA; [40] and the abagen toolbox [62] based on recommendations outlined by Arnatkeviciute et al. [6] Pearson correlations compared regional atrophy rates to gene expression patterns for each of the seven cell classes. These correlations were tested against spatial autocorrelation-preserving null models [4, 64].

We also compared the atrophy rate pattern to maps of different neurotransmitter receptor and transporter densities derived from PET imaging of more than 1,200 total individuals [63]. These 19 receptors and transporters spanning nine neurotransmitter systems included: acetylcholine (*α*_4_, *β*_2_, M_1_, VAChT), cannabinoid (CB_1_), dopamine (D_1_, D_2_, DAT), GABA (GABA_*A/BZ*_, histamine (H_3_), glutamate (mGluR_5_, NMDA), norepinephrine (NET), opioid (MOR), and serotonin (5-HT1_*A*_, 5-HT1_*B*_, 5-HT2_*A*_, 5-HT_4_, 5-HT_6_, 5-HTT). Volumetric tracer maps were obtained from https://github.com/netneurolab/hansen_receptors, parcellated to the Cammoun atlas and individually *z*-scored using the neuromaps toolbox [63]. Tracers for which multiple density maps were available were combined using weighted averaging. Pearson correlations compared regional atrophy rates to receptor/transporter expression and were tested against spatial autocorrelation-preserving null models [4, 64].

Lastly, we tested if there was asymmetry in cortical thinning between hemispheres, both at baseline and with disease progression. For each PD patient, mean cortical thickness was calculated in each hemisphere and an asymmetry index was computed based on the following formula:

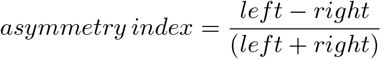

with more positive scores reflecting greater left dominance and more negative scores denoting greater right dominance. Asymmetry index scores were submitted to a linear mixed effects model to examine the change in cortical thickness asymmetry across follow-up visits.

### Structural and functional networks

Structural and functional data from *n* = 70 healthy young adults (25.3 *±* 4.9 years; 16 females) were obtained from the publicly available Lausanne dataset [36]. Imaging was acquired in a 3T MRI scanner (Trio, Siemens Medical) with a 32-channel head coil. Acquisition parameters and preprocessing steps are described in greater detail elsewhere [**?**].

Diffusion spectrum imaging (128 diffusion-weighted volumes and a single *b*_0_ volume, maximum *b*-value = 8,000 s/mm^2^, voxel size = 2.2 × 2.2 × 3.0 mm) was analyzed with deterministic streamline tractography to generate individual structural connectivity networks. These networks were parcellated according to the Cammoun atlas. A binary group-consensus structural network was created using an approach that preserved the density and edge-length distributions of individual connectomes [12]. A weighted structural network was then generated by weighting edges in the binary network by the log-transform of non-zero fiber density, scaled to values between 0 and 1.

Resting-state MRI data was acquired with a gradient echo EPI sequence sensitive to blood-oxygen-level-dependent (BOLD) contrast (3.3 mm in-plane resolution and slice thickness with 0.3 mm gap, TR = 1920 ms, 280 volumes). Time series data were parcellated according to the Cammoun atlas. Pairwise Pearson correlations were used to estimate individual functional connectivity networks. A group-average functional network was constructed as the mean pairwise connectivity across individuals. One individual did not have fMRI data and therefore the functional network was derived from *n* = 69 participants.

### Network spreading analysis

Next, we related the atrophy progression pattern to brain connectivity using a network spreading approach [89]. For each region (or node), we calculated a neighbourhood atrophy rate. This score described the mean atrophy rate across anatomically connected neighbours weighted by the strength of connectivity. Connectivity was defined using structural and functional reference networks derived from a separate cohort of healthy participants (see *Structural and functional networks*). Neighbourhood atrophy rate was calculated as follows:

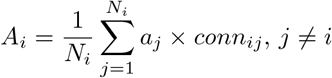

where *A*_*i*_ is the average neighbour atrophy rate of node *i, a*_*j*_ is the atrophy rate of the *j*-th neighbour of node *i, conn*_*ij*_ is the strength of connection between nodes *i* and *j*, and *N*_*i*_ is the total number of neighbours that are structurally connected to node *i* (i.e., node degree). Neighbour atrophy rate is made independent from node degree by normalization with term *N*_*i*_. For a given node, a neighbour was defined as another structurally connected node. Self-connections of a node and itself were excluded (*j* ≠ *i*). Connectivity (*conn*_*ij*_) was defined either structurally or functionally in separate analyses. Pearson correlations tested the relationship between the atrophy rate of each node with that of its neighbourhood and compared against spatial autocorrelation-preserving null models [4, 64]. We also examined the importance of structural and functional connectivity to node-neighbour correlations by comparing connected versus not-connected neighbours using Fisher’s *z* tests.

### Gene expression

To understand the gene expression profiles underpinning cortical atrophy progression patterns, regional gene expression data were obtained from the AHBA [40] dataset and processed with the abagen toolbox [64]. The AHBA is a comprehensive atlas of microarray data from six post-mortem brains (1 female, mean age = 42.5 *±* 13.4 years). Microarray probes were first reannotated following previous recommendations [6]. Only those probes with a signal-to-noise ratio relative to background of greater than 50% were retained. When multiple probes indexed expression of the same gene, the one with the most consistent pattern of regional variation across donors was selected. This procedure resulted in 15,633 total genes being retained. Samples were assigned to parcels according to the Cammoun atlas. To increase spatial coverage, tissue samples were mirrored bilaterally across the left and right hemisphere [74, 83, 87]. When a sample was not found directly within a parcel, the nearest sample (up to a 2 mm distance) was selected. If no samples were found within 2 mm of a parcel, the sample closest to the centroid of the empty parcel across all donors was selected. This sample-to-region matching was restricted to each hemisphere and within gross structural divisions (i.e., cortex, subcortex/brainstem, and cerebellum) to minimize the potential for misassignment. Samples without an assignment were discarded. Expression values were normalized across genes using a scaled robust sigmoid function [32] and rescaled to the unit interval. Microarray samples belonging to the same parcels were aggregated by computing the mean expression across samples for the individual parcels, for each donor. Finally, regional expression profiles were averaged across donors to construct a region × gene expression matrix that was submitted to a PLS regression.

### Gene set enrichment analysis

We used PLS analysis to uncover the gene expression profiles that predicted atrophy progression in PD. Latent variables that explained the maximum covariance between predictor matrix *X* (68 regions × 15,633 genes) and matrix *Y* (68 region × 1 atrophy rate) were identified. To determine the statistical significance of components (i.e., that the variance explained was greater than expected by a null), empirical variance was tested against the variance observed in 10,000 spatial autocorrelation-preserving null models. The contribution of genes to each component was measured using bootstrap resampling. Rows in *X* and *Y* matrices were randomly shuffled and PLS regression was repeated 5,000 times to generate a null distribution and estimate standard errors for each gene. Bootstrap ratios were calculated as the ratio of each gene expression weight to its bootstrapped-estimated standard error. Genes with large bootstrap ratios contributed greatly to the latent variable and were more reliable. Genes lists were ordered according to their bootstrap ratios and this ranked list was entered into a gene set enrichment analysis (GSEA). GSEA, performed on the WebGestalt platform (http://www.webgestalt.org; [54]) with the Gene Ontology knowledge base (http://geneontology.org), was used to explore the biological processes enriched in the gene expression profiles associated with atrophy rates in PD. This analysis tested whether the most positively and negatively weighted genes in the ranked list occurred more frequently than expected by chance [94]. The minimum and maximum number of genes for enrichment was set to 3 and 2,000, respectively. Results were FDR-corrected for multiple comparisons using 1,000 random permutations. Only the top 10 most significant positively and negatively weighted terms were interpreted.

## RESULTS

### Participants

After completing quality control of the cortical thickness maps, the final sample was comprised of *n* = 160 patients contributing 478 scans (151 at baseline, 121 at year-1, 114 at year-2, and 82 at year-4) and *n* = 137 healthy controls at baseline. The two groups were similar to one another at baseline in terms of age (PD = 61.1 *±* 9.8 years, HC = 59.9 *±* 11.4 years; *t*(286) = 0.97, *p* = 0.37) and sex (*χ*^2^ = 0.25, *p* = 0.61). Table 1 displays demographic and clinical measure scores for PD patients included in the present study. Linear mixed effects models of clinical progression in PD patients revealed a significant effect of time for measures of disease stage (i.e., Hoehn Yahr score), motor symptoms (i.e., MDS-UPDRS-II, MDS-UPDRS-III, tremor, rigidity), non-motor symptoms (e.g., MDS-UDPRS-I, Scales for Outcomes in Parkinson’s Disease-Autonomic, REM sleep behavior disorder, phonemic fluency), and CSF protein levels (amyloid beta, phosphorylated and total tau, neurofilament light chain [NfL]). Generally, these results reflected a worsening of clinical symptoms over time except for phonemic fluency, which improved across visits. Whereas amyloid beta, phosphorylated tau, and total tau levels in CSF decreased over time, NfL levels increased.

### Longitudinal changes in cortical atrophy

Linear mixed effects models of whole-brain cortical thickness revealed a significant effect of time (*t* = 4.955, *p* < 0.001) reflecting increased atrophy across visits. Region-wise analysis of cortical thickness showed a largely posterior parietal, temporal, and superior frontal pattern of atrophy progression (Fig. 1a). No regions showed statistically significant increases in cortical thickness over time (Fig. 1b). The spatial pattern of atrophy progression observed was consistent across parcellation resolutions (Fig. S1). Overall, we observed a widespread pattern of longitudinal cortical atrophy progression in PD patients.

**Figure 1:**
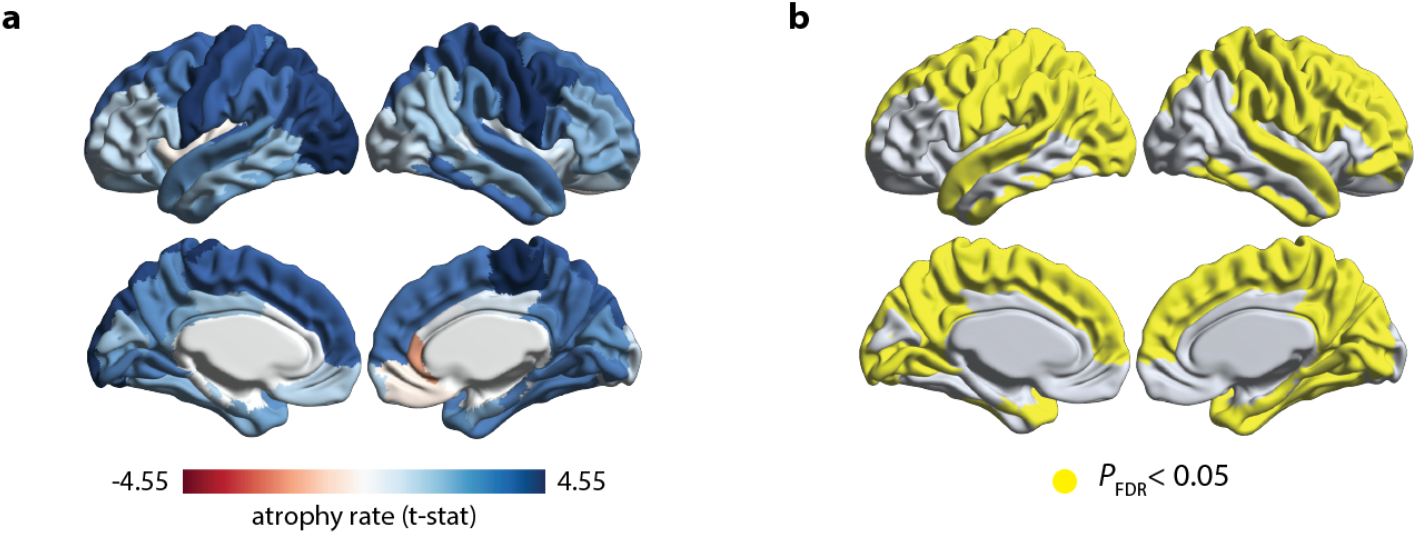
Pattern of cortical atrophy progression in Parkinson’s disease. **(a)** Rates of cortical thickness changes (*t*-statistic of ‘time’ effect) for linear mixed effect models fit to longitudinal MRI data in Parkinson’s disease patients. Bluer regions denote greater atrophy progression over 4-year follow-up. **(b)** Cortical regions demonstrating significant ‘time’ effect after FDR-correction for multiple comparisons (*p*_*FDR*_ < 0.05).

### Atrophy-clinical progression

PLS was used to relate changes in atrophy and clinical progression in PD. We identified the first latent variable that accounted for 49.1% (*p* < 0.001) of the covariance between these measures. Regional brain weights associated with this latent variable were positively correlated with atrophy rates (*r* = 0.309, *p*_*spin*_ = 0.028; Fig. 2b), such that brain regions with more positive weighting on the latent variable also demonstrated greater atrophy rates (Fig. 2a). Of the 25 longitudinal clinical measures included in the analysis, five measures significantly contributed to the latent variable (Fig. 2c), namely the Benton Judgement of Line Orientation test (Benton JLO), the Symbol-Digit Modalities test, the Questionnaire for Impulsive-Compulsive Disorders in Parkinson’s Disease-Rating Scale (QUIP), the Scales for Outcomes in Parkinson’s Disease-Autonomic (SCOPA-AUT), and CSF levels of neurofilament light chain (NfL). In other words, PD-related cortical atrophy progression co-vary most with changes in visuospatial perception, psychomotor slowing, impulsive and compulsive behaviours, autonomic dysfunction, and CSF levels of NfL.

**Figure 2:**
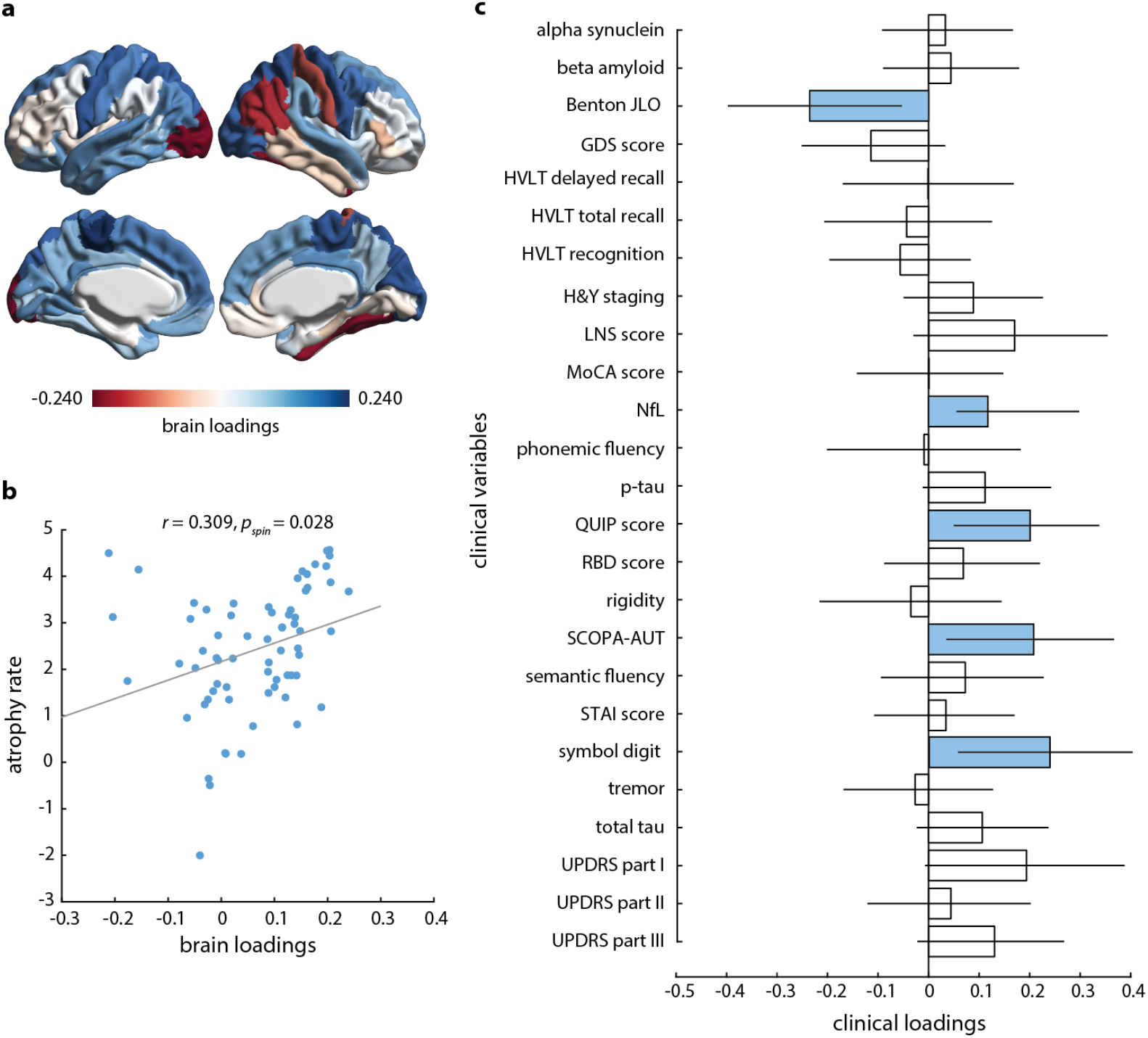
Atrophy-clinical progression relationship. **(a)** Brain loadings (i.e., bootstrap ratios) associated with the first significant latent variable from a partial least squares analysis between individual patient atrophy rates and clinical variable progression. Bluer regions represent more positive contributions to the expression of the atrophyclinical latent variable. **(b)** Scatter plot of the correlation between brain loadings and atrophy rates. Regions with more positive loadings on the latent variable corresponded to regions with greater atrophy rates. **(c)** Clinical variable loadings associated with the first significant latent variable. Blue boxes indicate clinical variables that significantly contributed to the latent variable. Error bars represent 95% confidence intervals. Details on each clinical variable can be found in Table 1.

### Cortical atrophy progression is distributed within discrete brain systems

Spatial correspondence analyses tested whether the pattern of atrophy progression in PD was pronounced in specific brain systems. Specifically, we examined the relationship between atrophy rates and the (i) Yeo intrinsic networks [109], (ii) von Economo cytoarchitectonic classes [84, 104], (iii) cell type distributions, and (iv) neurotransmitter receptor and transporter densities [63]. Atrophy rates were relatively greater in the sensorimotor network (*mean* = 3.636, *p*_*spin*_ = 0.018; Fig. 3a) and relatively lesser in the ventral attention network (*mean* = 1.516, *p*_*spin*_ = 0.014; Fig. 3a). We also noted relatively greater atrophy rates within primary motor cortex (*mean* = 4.309, *p*_*spin*_ = 0.022; Fig. 3b). The atrophy progression pattern was negatively correlated with astrocyte cell distribution (*r* = -0.451, *p*_*spin*_ = 0.019; Fig. 3c). Regions with greater expression of astrocytes corresponded with relatively lower atrophy rates. Finally, atrophy progression was negatively correlated with the distribution of several receptors and transporters, including: 5HTT (*r* = -0.361, *p*_*spin*_ = 0.024), CB_1_ (*r* = -0.460, *p*_*spin*_ = 0.040), D_1_ (*r* = -0.604, *p*_*spin*_ <0.001), DAT (*r* = -0.470, *p*_*spin*_ = 0.001), H_3_ (*r* = -0.504, *p*_*spin*_ = 0.005), mGluR_5_ (*r* = -0.393, *p*_*spin*_ = 0.011), and MOR (*r* = -0.551, *p*_*spin*_ = 0.009). Atrophy progression was negatively correlated with each of these neurotransmitter systems (Fig. 3d), suggesting that regions with lower expression of these neuroreceptors displayed higher atrophy rates. Finally, cortical asymmetry was not found between left and right hemispheres at baseline (*t* = - 0.764, *p* = 0.445) and did not progress across visits (*t* = -1.00, *p* = 0.317). Collectively, we found that the pattern of atrophy progression in PD is spatially distributed within discrete brain systems and reflects underlying biology including astrocyte cellular architecture and neurotransmitter systems.

**Figure 3:**
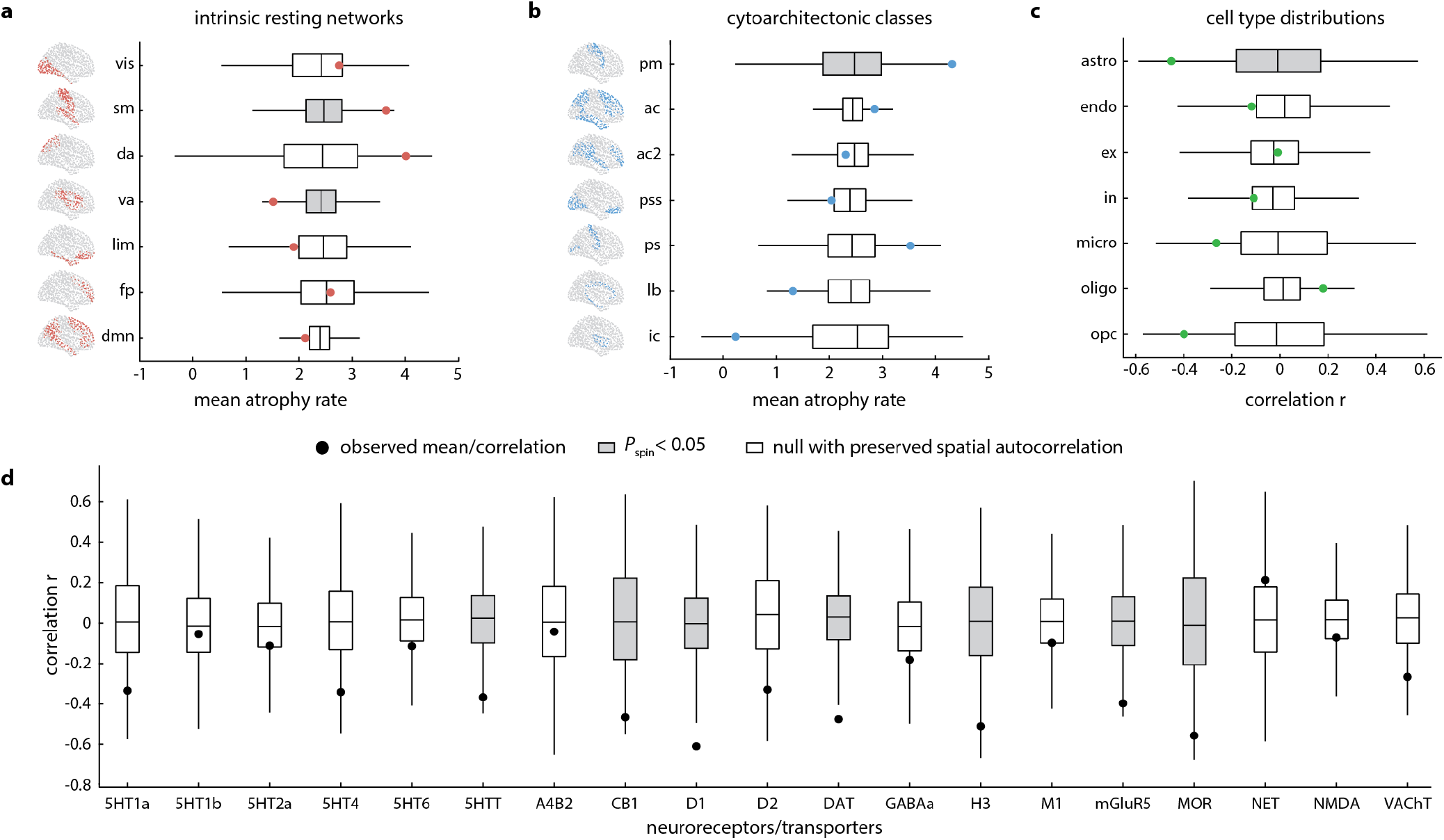
Distribution of cortical atrophy progression within different brain systems. **(a)** Box plots of mean atrophy rates within intrinsic resting networks (vis = visual, sm = sensorimotor, da = dorsal attention, va = ventral attention, lim = limbic, fp = frontoparietal, dmn = default mode) and **(b)** cytoarchitectonic classes (pm = primary motor, ac/ac2 = association 1/2, pss = primary/secondary sensory, ps = primary sensory, lb = limbic, ic = insular. **(c)** Box plot of correlations between regional atrophy rates and cell type distributions (astro = astrocytes, endo = endothelial cells, ex/in = excitatory and inhibitory neurons, micro = microglia, oligo = oligodendrocytes, opc = oligodendrocyte precursors). **(d)** Boxplot of correlations between regional atrophy rates and 19 different neurochemical maps: acetylcholine (*α*_4_,*B*_2_, M_1_, VAChT), cannabinoid (CB_1_), dopamine (D_1_, D_2_, DAT), GABA (GABA_*A/BZ*_, histamine (H_3_), glutamate (mGluR_5_, NMDA), norepinephrine (NET), opioid (MOR), and serotonin (5-HT1_*A*_, 5-HT1_*B*_, HT2_*A*_, 5-HT_4_, 5-HT_6_, 5-HTT). Grey boxes denote significant means or correlations when tested against spatial autocorrelation-preserving null models (*p*_*spin*_ < 0.05).

### Structural and functional network connectivity constrains cortical atrophy progression

Using a network spreading approach (Fig. 4a; [89]), we tested the relationship between the atrophy rate measured in each node and the mean atrophy rate across all structurally and functionally connected neighbours. We found positive correlations between node and neighbourhood atrophy rates (Fig. 4b), suggesting regions with greater atrophy rates are themselves connected to neighbourhoods with overall greater atrophy rates. This was the case whether connectivity was defined structurally (*r* = 0.470, *p*_*spin*_ = 0.018; Fig. 4b left) or functionally (*r* = 0.425, *p*_*spin*_ = 0.049; Fig. 4b right). Importantly, these results were robust to different parcellation resolutions and when tested against spatial autocorrelation-preserving null models (Fig. S3).

**Figure 4:**
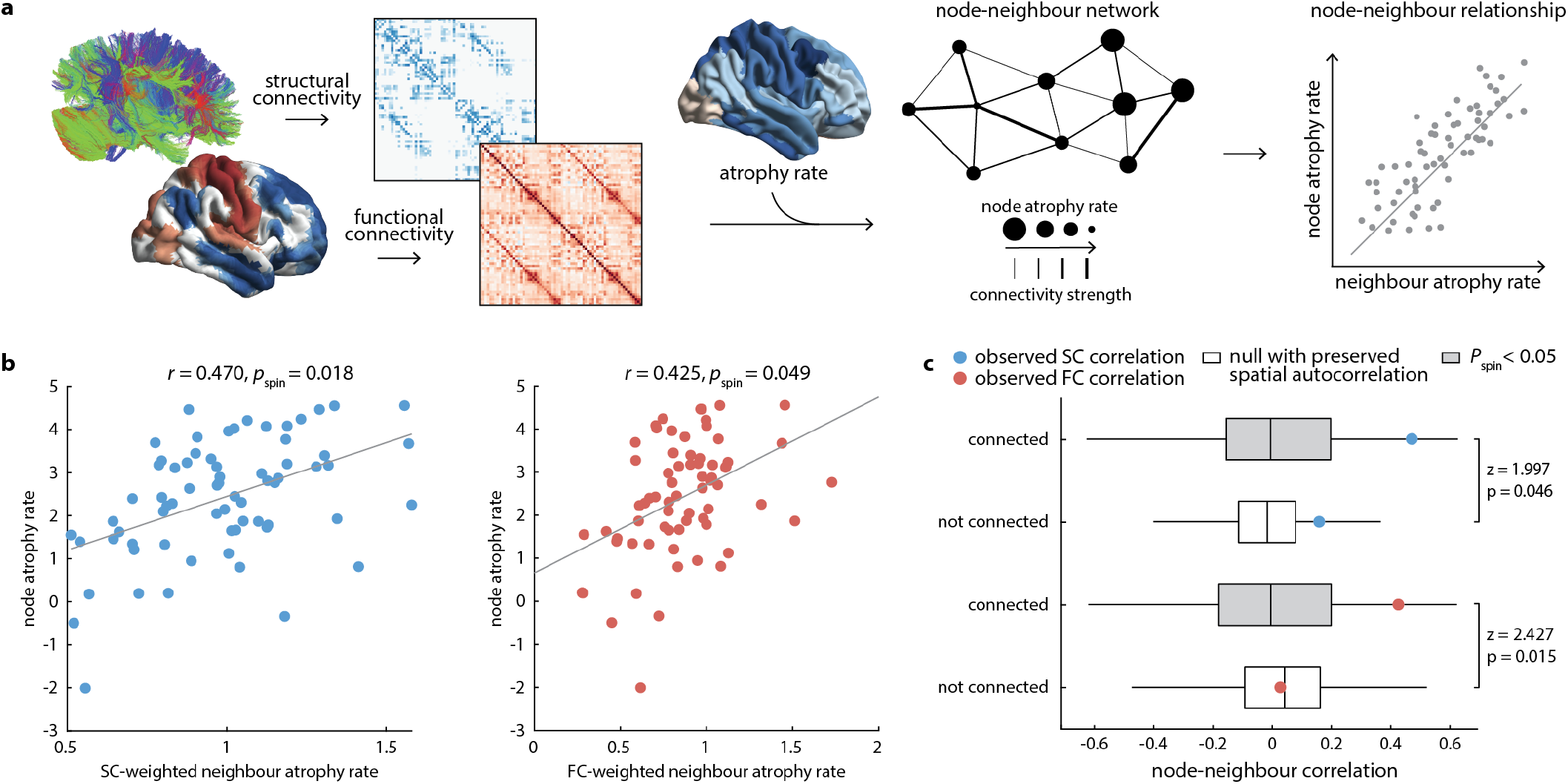
Network spreading models of cortical atrophy progression. **(a)** Schematic of network spreading model. Reference structural and functional connectivity networks derived from a separate cohort of healthy subjects and the atrophy rate map generated from Parkinson’s disease patients were used to compute neighbourhood atrophy rates—the average atrophy rate across anatomically connected neighbours weighted by structural or functional connectivity strength—for each region or node. **(b)** Scatter plots of the relationships between node versus neighbourhood atrophy rates for structurally (left) and functionally (right) connected neighbours. **(c)** Box plots comparing correlation coefficients (*r*) for node-neighbourhood relationships from networks composed of connected versus not-connected neighbours. Grey boxes indicate statistical significance against spatial autocorrelation-preserving null models (*p*_*spin*_ < 0.05). Fisher’s *z* tests compared connected versus not-connected *r* values.

We also considered node-neighbour relationships when networks were composed of connected versus not-connected neighbours. Fisher’s *z* tests revealed that the magnitudes of correlations with connected neighbours were significantly greater than those for not-connected neighbours, whether defined structurally (*z* = 1.997, *p* = 0.046) or functionally (*z* = 2.427, *p* = 0.015; Fig. 4c). Node-neighbour correlations for structurally and functionally connected neighbours were significantly greater than not-connected neighbours across parcellation resolutions, except for functional connectivity at the highest spatial scale (i.e., 1000 regions; Fig. S3) that did not reach statistical significance. Taken together, we demonstrate the influence of network architecture in shaping the pattern of cortical atrophy progression in PD.

### Genes related to atrophy progression are enriched for mitochondrial and metabolic processes

PLS analysis was used to relate regional gene expression to the atrophy progression pattern in PD. The first (i.e., PLS1) and second (i.e., PLS2) significant latent variables explained 30.4% (*p*_*spin*_ = 0.027) and 26.8% (*p*_*spin*_ = 0.028) of the observed co-variance, respectively. Atrophy rates were positively correlated with brain weights that contributed to PLS1 (*r* = 0.540, *p*_*spin*_ = 0.012; Fig. 5a-b) and PLS2 (*r* = 0.519, *p*_*spin*_ < 0.001; Fig. 5d-e). In regions with greater atrophy rates, those genes that positively weighted on the latent variables were more expressed whereas those genes that negatively weighted on the latent variables were less expressed. Next, genes were ranked in order of their bootstrap ratios separately for each latent variable and examined for their biological relevance using GSEA. The most positively weighted genes (i.e., genes more expressed in regions with greater atrophy rates) were enriched for processes related mitochondrial and metabolic function (see Table S1 and S2). For genes positively associated with PLS1, the most enriched terms included “mitochondrial RNA metabolic process” (*normalized enrichment score* = 1.951, *p*_*FDR*_ = 0.037; Fig. 5c) and “mitochondrial gene expression” (*normalized enrichment score* = 1.889, *p*_*FDR*_ = 0.046; Fig. 5c). For genes positively related to PLS2, regions with greater atrophy rates had greater expression of genes involved in the “NADH dehydrogenase complex assembly” (*normalized enrichment score* = 1.967, *p*_*FDR*_ = 0.032; Fig. 5f) and “mitochondrial gene expression” (*normalized enrichment score* = 1.949, *p*_*FDR*_ = 0.024; Fig. 5f). Negatively weighted genes associated with either latent variable was not found to be significantly enriched for specific biological processes. In summary, genes more expressed in regions with greater atrophy rates in PD are associated with mitochondria and energy metabolism.

**Figure 5:**
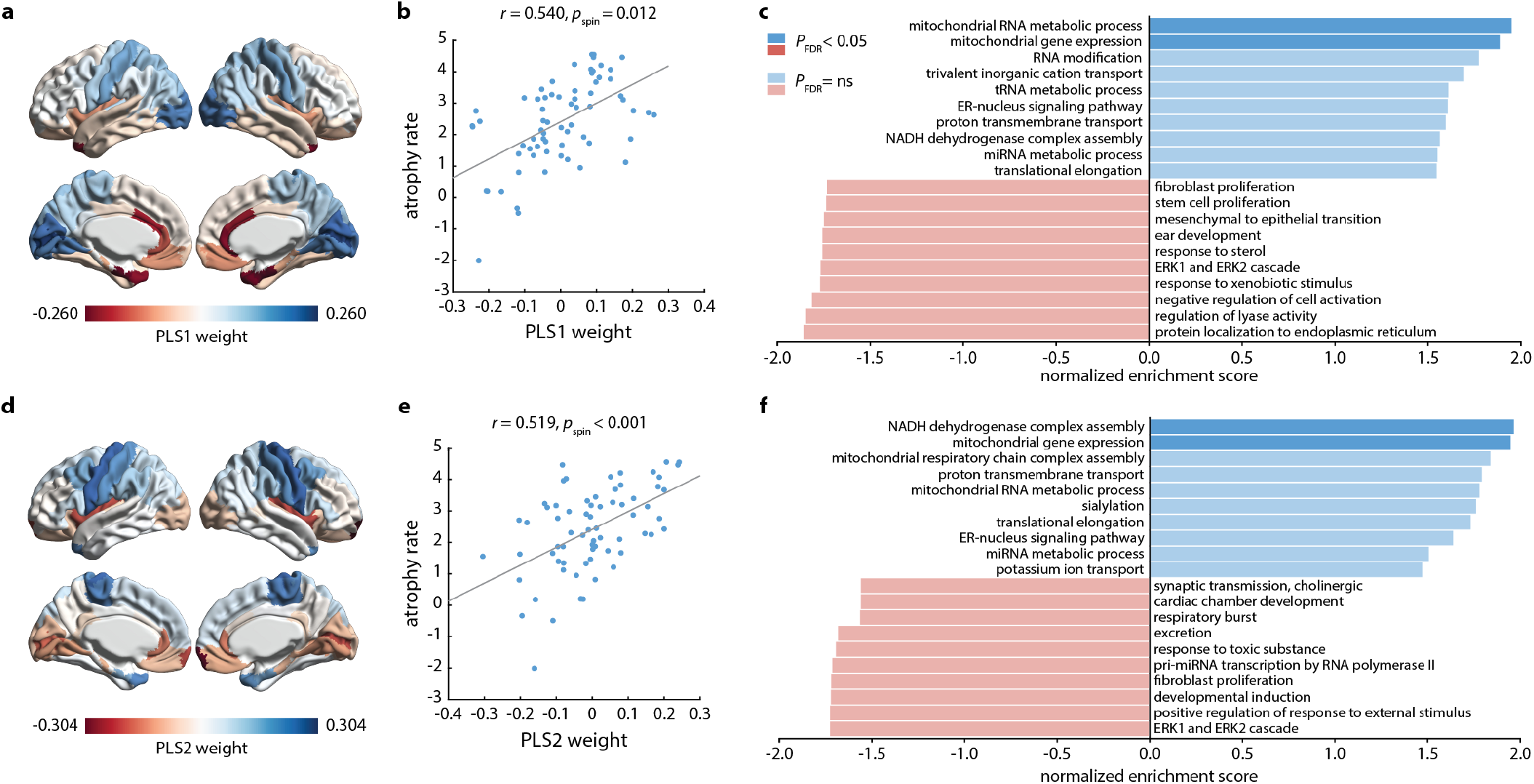
Relationship between regional atrophy rates and gene expression profiles. Brain loadings for the **(a)** first and **(d)** second significant latent variables (i.e., PLS1 and PLS2, respectively) from a partial least squares analysis relating regional atrophy rates to gene expression profiles derived from the Allen Human Brain Atlas. Scatter plots of the relationship between **(b)** PLS1 and **(e)** PLS2 brain loadings and regional atrophy rates. Brain regions that contributed more positively to their respective latent variables and expressed more positively weighted genes also had greater atrophy rates. Enriched biological processes genes expression profiles associated with **(c)** PLS1 and **(f)** PLS2. More positive enrichment scores were associated with patterns of greater atrophy rates. Darker blue and red bars indicate statistically significant enrichment scores (*p*_*FDR*_ < 0.05).

## DISCUSSION

The present study aimed to characterize and better understand the progression of cortical atrophy in early PD. We found that cortical atrophy progresses within mostly posterior and temporal regions as well as superior frontal cortex over 4 years. We also found the progression of cortical atrophy occurred in association with the clinical progression of motor and non-motor features, and CSF tau and NfL levels. Moreover, we demonstrated that cortical atrophy progression in PD is underpinned by specific connectivity and gene expression features. Specifically, we showed that network architecture significantly constrained the progression of cortical atrophy and that this pattern overlapped with motor network and cytoarchitectonic classes, astrocyte cellular composition, and several neurochemical distributions. In addition, cortical atrophy progression was related to gene expression profiles enriched for mitochondrial and metabolic functions. Taken together, our results shed light on the global network and local features that shape disease progression in PD.

### Cortical atrophy and clinical outcome progression

We observed progression of cortical atrophy in PD within posterior, temporal, and superior frontal cortices. Cortical thickness is thought to represent the volume of neuropil—the axons, dendrites, and synapses within cortical columns—and therefore cortical thinning likely reflects the loss of neuropil and synaptic density [102]. The observed pattern was largely consistent with that found in case-control [51, 73, 98, 99] and longitudinal [2, 60, 96, 97, 107] studies of PD. Along with regional atrophy progression, worsening clinical outcomes were also found in several motor (i.e., HY stage, UPDRS-II, UPDRS-III), non-motor (i.e., UPDRS-I, RBDSQ, SCOPA-AUT), and CSF biomarker (i.e., beta-amyloid, p-tau, t-tau, NfL) measures. Worsening motor symptoms and autonomic dysfunction are expected with advancing disease [26, 112]. Although measures of verbal fluency were found to improve with time, this likely reflects practice or medication effects.

A multivariate analysis identified a single latent variable that best explained the covariance between the progressions of cortical atrophy and clinical outcomes in this PD cohort. The clinical changes that significantly contributed to this latent variable were largely non-motor (i.e., visualspatial perception, psychomotor slowing, compulsive-impulsive behaviours, autonomic dysfunction) and CSF biomarker (i.e., NfL) measures. Notably, changes in motor symptom measures did not correlate with atrophy progression in this analysis despite a clear progression of these scores in our linear mixed effects models. This might owe to the fact that our analysis was restricted to atrophy within the cortex, whereas motor symptoms are almost entirely due to subcortical effects of the disease, especially loss of dopamine signalling in the putamen [49] Examining the atrophyclinical progression relationship in subcortical regions might reveal an association with motor symptoms. Indeed, a previous PLS analysis performed on whole-brain atrophy and clinical data in *de novo* PD patients from the PPMI at baseline uncovered a clinical phenotype that included both motor and non-motor features associated with diffuse tissue loss [110]. Further, although *de novo* PD patients were not yet treated with medications at the baseline visit, they are prescribed medications increasingly, which could improve their motor symptoms even as the disease worsens.

We observed longitudinal changes in several CSF biomarkers, including decreases in tau (total tau and p-tau) and beta-amyloid but increases in NfL levels. Although both CSF tau and beta-amyloid are typically markers of Alzheimer’s disease, many PD patients demonstrate co-pathologies that correlate with cognitive decline and reduced survival [43]. Longitudinal modeling of these CSF biomarkers in PD is limited, however a recent study performed on the PPMI cohort found lower levels of p-tau and beta-amyloid in PD patients compared to healthy controls [42], a trend with which our results are in line. Lastly, there was a noted elevation in NfL concentrations in CSF over time. Neurofilaments are cytoskeletal components of myelinated axons that are released following axonal degeneration [70]. Of the several subclasses of neurofilaments, NfL is the most thoroughly investigated as a biomarker of PD progression [33, 46]. Studies have shown that NfL levels in blood and CSF are significantly elevated in PD patients compared to healthy controls, and baseline NfL levels are predictive of motor and cognitive progression in PD [1, 9, 68, 108]. Notably, post-mortem measures of NfL immunoreactivity correlated with MRI-derived cortical thinning in PD patients [30]. Here we add to this literature by showing that progressive increases in NfL covary with regional cortical atrophy rates.

### Distribution of cortical atrophy progression

The atrophy progression pattern was found to be localized to specific brain systems. Relatively greater atrophy rates were observed in sensorimotor network and primary motor cortex, whereas relatively lower atrophy rates were found in ventral attention network. Motor and sensory areas of the cortex are primarily connected to the putamen, which is severely dopamine-depleted in PD [48]. Previous work has shown that cortical thinning is determined by connectivity to a subcortical “disease reservoir” in PD, suggesting a potential pathway by which PD pathology originating in subcortex might dominate specific regions or networks of neocortex [107].

Additionally, regions with higher expression of astrocytes corresponded to regions with lower atrophy rates. Whether the role of astrocytes in PD is deleterious or protective has yet to be settled [91]. For example, some evidence suggests that alpha-synuclein aggregates readily exchanged between neurons and astrocytes produce an inflammatory response and lead to neuronal death if transferred from astrocytes to neurons [20, 41]. In contrast, others have shown that astrocytes may take up and degrade misfolded alpha-synuclein more efficiently than neurons [55]. This neuroprotective function seems in line with our observation of relatively less atrophy progression in astrocyte-rich regions, although more clarification is needed.

Finally, regions with higher atrophy rates overlapped areas with relatively lower distribution of several neurochemical systems including serotonin (5-HTT), cannabinoid (CB_1_), dopamine (D_1_, DAT), histamine (H_3_), glutamate (mGluR_5_), and opioid (MOR). The involvement of these various transmitters and receptors in PD pathology has been described previously and mapped to largely frontal and striatal regions [5, 13, 18, 75, 81]. Indeed, visual inspection of the cortical topography of these neurochemical maps (see Fig. S2) reveals their concentration in ventral frontal and limbic cortices, where we observed the lowest atrophy rates. Although we saw a negative association between atrophy rates and D_1_/DAT concentrations (i.e., greater atrophy in regions with lower concentrations of dopamine receptor and transporter), note that our analysis was restricted to the cortical surface whereas dopamine deficiency caused by PD occurs primarily in basal ganglia [49]. Nonetheless, these results reveal interesting associations between PD-related cortical atrophy and neurochemical mechanisms that warrant further exploration.

### Network architecture shapes cortical atrophy progression

There is increasing evidence of a network spreading process of atrophy in multiple neurodegenerative diseases and psychiatric disorders, including PD, Alzheimer’s disease, frontotemporal dementia, amyotrophic lateral sclerosis, and schizophrenia [29, 67, 71, 76, 80, 85, 88, 89, 96, 107, 115]. Although these diseases have distinct aetiologies, the fact that neurodegeneration follows large-scale brain networks raises the possibility of similar pathophysiological mechanisms. One hypothesis posits that neurodegenerative diseases spread through the cell-to-cell propagation of toxic agents in a prion-like manner [45, 105]. In PD, that agent is a misfolded isoform of alpha-synuclein spreading through the connectome [31, 45, 105].

We found evidence that cortical atrophy progression in PD is shaped by structural and functional networks. Regions with greater atrophy rates were more likely to be connected to regions with greater atrophy rates themselves. Intrinsic anatomical and functional connectivity was important for this model, as networks composed of not-connected nodes disrupted this region-neighbourhood relationship. Critically, we ensured our results were independent of spatial autocorrelation and were replicable across spatial scales. These findings replicate and extend previous work in support of a network spreading model in both prodromal [78] and incidental PD [10, 71, 80, 96, 107, 111]. Zeighami et al.[111] first showed in /emphde novo PD patients that brain atrophy measured with deformation-based morphometry was associated with connectivity to the substantia nigra, a disease epicenter in PD. In a more recent study, Tremblay et al. [96] found that structural and functional connectivity continued to constrain deformation-based atrophy progression in PD patients followed longitudinally. Similarly, Yau et al. [107] reported that cortical thinning in /emphde novo PD over a 1-year period was correlated with connectivity to a subcortical “disease reservoir”. We build on these findings here by demonstrating the role of connectivity in the progression of cortical atrophy over a longer 4-year duration.

Alternatively, other pathogenic mechanisms would also follow the connectome. Connected areas tend to be similar in terms of cellular composition [106], neuroreceptor profiles [38], and gene expression [7]. Thus, a pathogenic process may target connected regions for reasons other than their connectivity. Highly connected hub regions in the brain tend to be targeted in almost all neurological diseases [22], possibly because of their precarious energy needs. These regions also tend to be inter-connected [100]. Indeed, we previously showed that atrophy patterns in the PPMI dataset preferentially target hub regions [111]. Nonetheless, many animal experiments confirm that pathogenic alpha-synuclein does propagate via anatomical connections [56, 65, 69]. Several human imaging studies, including this one, also support the model [59, 71, 80, 96, 107, 111].

### Atrophy progression-related genes are enriched for mitochondrial and metabolic function

Cortical atrophy progression was associated with gene expression profiles enriched for mitochondrial and metabolic function. Mitochondrial dysfunction was first implicated in PD based on the observation that exposure to the drug 1-methyl-4-phenyl-1,2,3,4-tetrahydropyridine (MPTP) induced the rapid onset of parkinsonism and degeneration in the substantia nigra of humans [52, 53]. Further inspection revealed this toxic effect was mediated by the inhibition of complex I of the mitochondrial electron transport chain. But even in cases of sporadic PD, reductions in complex I activity within the substantia nigra as well as in the cortex and periphery have been reported [34, 72, 93]. Interestingly, we identified NADH dehydrogenase complex as one of the most enriched terms in our GSEA. Specific genetic mutations in PD-related genes such as PINK and Parkin have also been implicated in defective mitochondrial function and regulation through impaired mitophagy of damaged mitochondria [14, 21, 58, 114]. Impaired mitochondria produce less cellular energy, but more reactive oxygen species linked to oxidative stress. The accumulation of dysfunctional mitochondria may contribute to regional vulnerability to PD pathology. Moreover, mitochondrial failure and accumulation of misfolded alphasynuclein may promote each other [82]. Our results add to this view by showing that cortical areas with more mitochondria-related gene expression are especially vulnerable to atrophy progression in PD. We also replicated findings from a previous study in prodromal PD patients that showed cortical atrophy-related genes were enriched for mitochondrial function [78].

## Limitations

We examined *de novo* PD patients followed longitudinally over a 4-year duration. This within-subject, multiple time point design afforded us both control of between-subject variability due to disease heterogeneity and a broader view of disease progression. Despite these advantages, this approach presents a few challenges. First, not all PD patients had MRI data available at all four visits due to either scans failing quality control for a subset of time points or because of attrition that is common among longitudinal patient studies. We therefore included only those individuals with data available at two or more time points and employed linear mixed effects modeling, which is robust to data with irregular timing and subject drop-out [11]. Next, although we examined PD patients over more frequent visits and across a longer span than most previous studies, still our results only reveal progression during a limited window of the entire PD course. Modeling repeated visits over a longer duration would allow for a more comprehensive estimation of disease trajectories. This approach might specifically better capture cortical changes and accompanying cognitive deficits presumed to be most pronounced later in the disease course.

Cortical thinning was used as a proxy for PD pathology. At present, direct measures of alpha-synuclein distribution in PD patients *in vivo* are not possible as there are no validated PET radiotracers for this protein. Such a tracer would allow for more accurate study of disease spread, as has been demonstrated in the case of beta-amyloid and tauopathies such as Alzheimer’s disease and frontotemporal dementia [39, 103]. Still, brain atrophy measured with MRI can arguably be used as an indirect surrogate of alpha-synuclein pathology in the brain.

The PPMI dataset consists of clinical and imaging data contributed from multiple centres and scanners, which might introduce unwanted heterogeneity [37]. However, the PPMI has strict and standardized protocols for data acquisition [61] and the CIVET pipeline used for analyzing cortical thickness in the present study was developed and validated to deal with such multi-centre datasets [23]. Further, we include scanning site as a covariate in our analyses of cortical atrophy and clinical progression.

## Conclusion

In summary, the present study reveals a distinct pattern of cortical atrophy progression across the early stage of PD. This pattern appears to be shaped by structural and functional network architecture as well as regional differences in cellular composition, neuroreceptor distribution, and gene expression. Notably, the gene expression profiles related to atrophy progression are associated with mitochondrial and metabolic functions. These results demonstrate how both global network and local vulnerability factors shape the progression of cortical atrophy in PD.

## Data Availability

Clinical and imaging data used in this study are part of the PPMI database and can be accessed at http://www.ppmi-info.org/data. All other datasets and analysis tools along with their sources are cited in the "Methods" section.

## ACKNOWLEDGEMENTS

PPMI—a public-private partnership—is funded by the Michael J. Fox Foundation for Parkinson’s Research and funding partners, including AbbVie, Avid, Biogen, Bristol-Myers Squibb, Covance, GE Healthcare, Genentech, GlaxoSmithKline, Lilly, Lundbeck, Merck, Meso Scale Discovery, Pfizer, Piramal, Sanofi Genzyme, Servier, Teva, and UCB. This research was supported by grants from the Michael J. Fox Foundation for Parkinson’s Research, the W. Garfield Weston Foundation, the Alzheimer’s Association, the Canadian Institutes of Health Research, and the Natural Sciences and Engineering Research Council of Canada to AD. AV acknowledges support from Frontiers de Rechèrche du Québec – Santé (FRQS). SR received a scholarship from the Canada Institutes of Health Research (CIHR).

