## Supplemental Materials for "Network connectivity and local transcriptomic vulnerability underpin cortical atrophy progression in Parkinson’s disease"

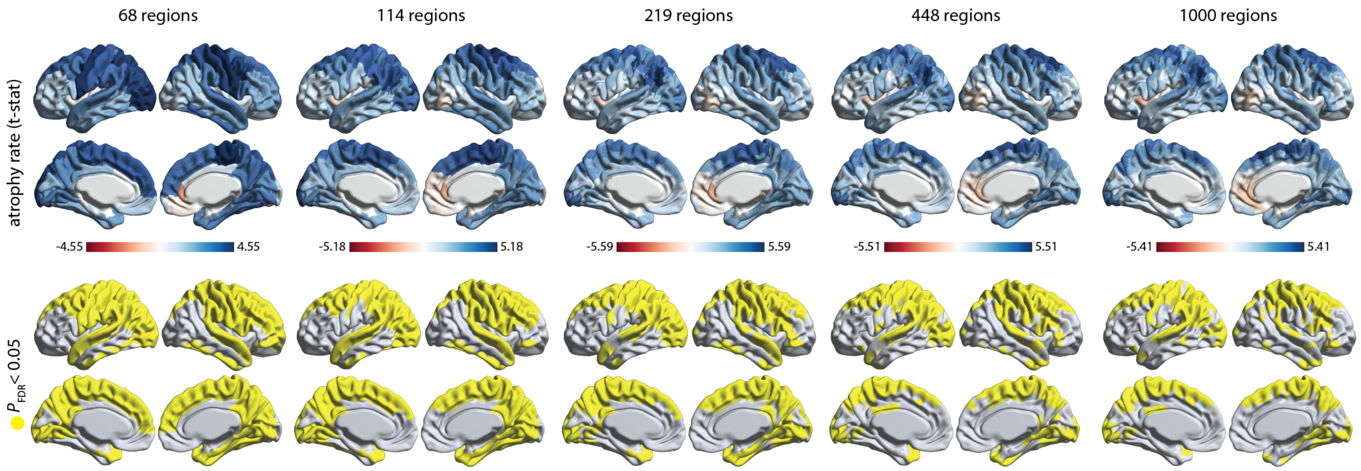


Figure S1: **Cortical atrophy rates across parcellation resolutions.** The cortical atrophy progression analysis was repeated across five progressively finer anatomical parcellations. *Top row:* Regional atrophy rates ($t$-statistic of 'time' effect from linear mixed effects model). *Bottom row:* Regions with significant atrophy rates after correction for multiple comparisons ($p$<0.05).

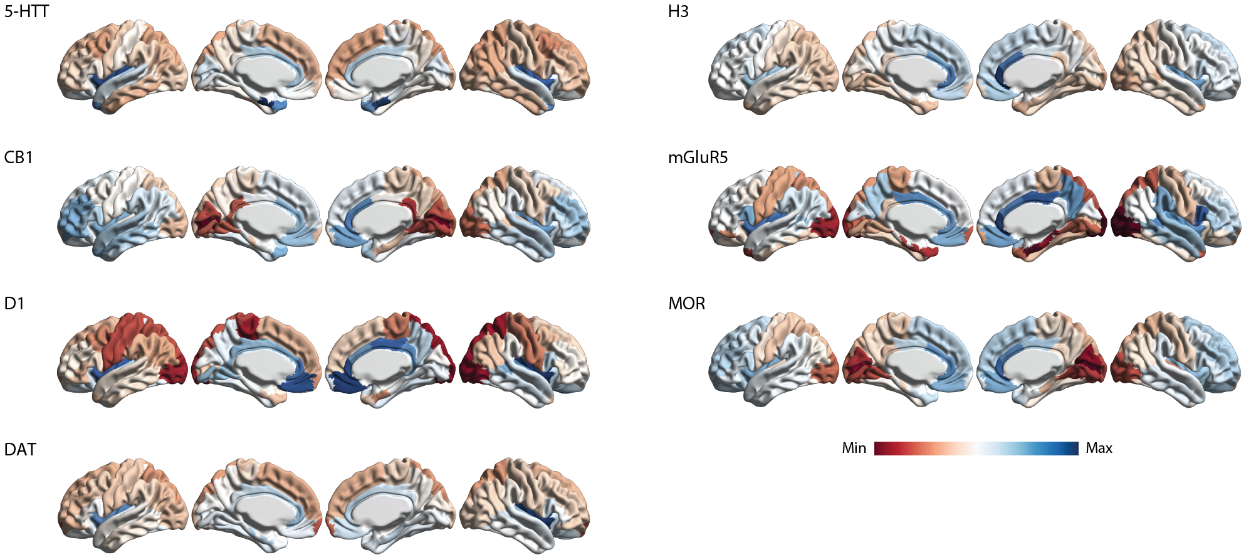


Figure S2: **Expression maps for neurotransmitter receptors and transporters on the cortical surface.** Surface maps of distributions for neuroreceptor systems that significantly correlated with regional atrophy rates. 5-HTT = serotonin; $CB_1$ = cannabinoid receptor; $D_1$ = dopamine $D_1$ receptor; DAT = dopamine transporter; $H_3$ = histamine; $mGluR_5$ = glutamate receptor; MOR = opioid receptor

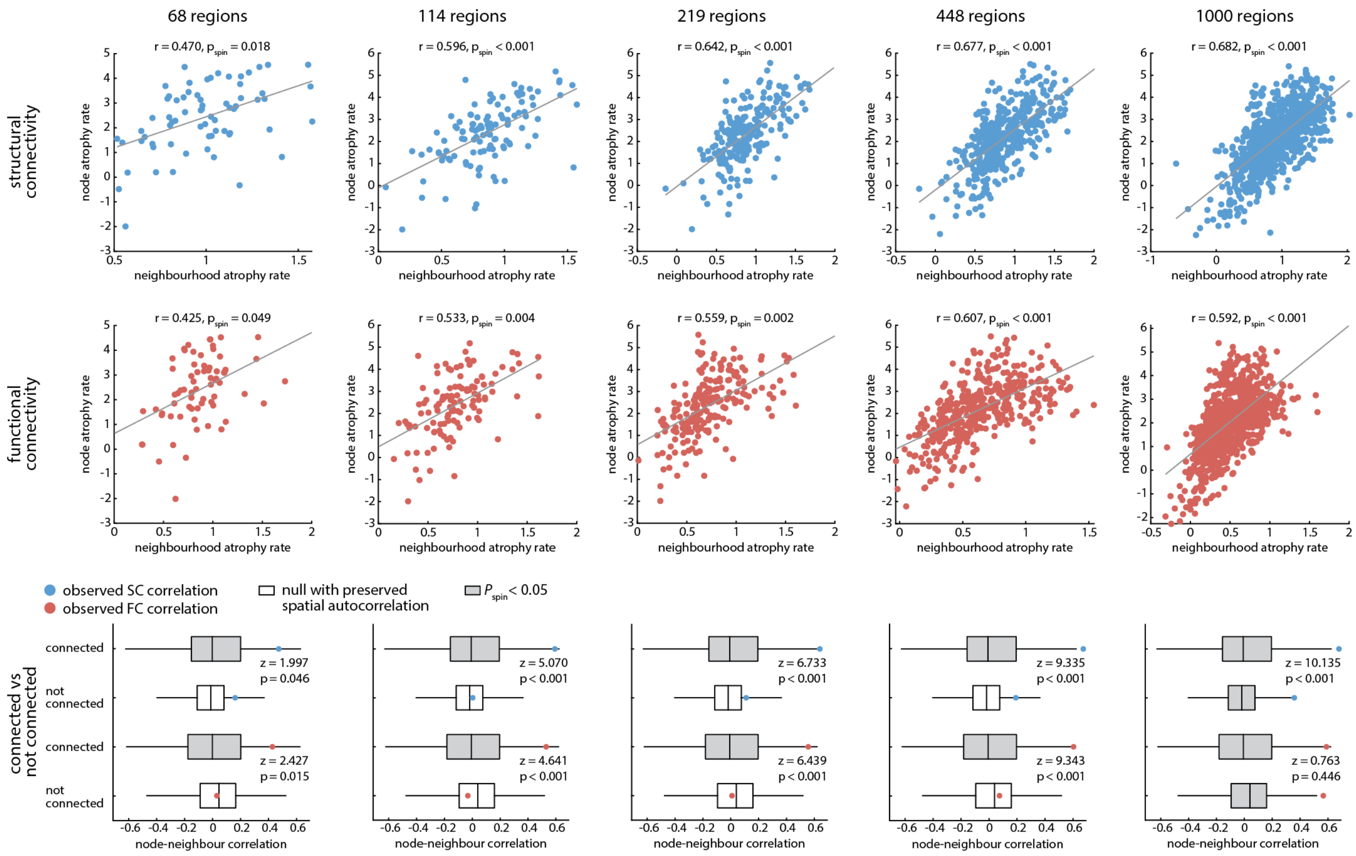


Figure S3: **Node-neighbourhood correlations across parcellation resolutions.** The network spreading analysis was repeated across five progressively finer anatomical parcellations for structurally- (*top*) and functionally- (*middle*) weighted networks. *P*-values were determined from 10,000 spatial autocorrelation-preserving null model tests. Correlation $r$ values for networks composed of connected versus not-connected nodes were compared using Fisher's $z$ score (*bottom*).

TABLE S1: **Summary of gene set enrichment analysis for the first significant latent variable (PLS1).**

| **Gene Set** | **Description** | **Size** | **Leading Edge Number** | **ES** | **NES** | ***P*-Value** | **FDR** |
|---|---|---|---|---|---|---|---|
| GO:0000959 | mitochondrial RNA metabolic process | 40 | 17 | 0.540 | 1.951 | <0.001 | 0.037 |
| GO:0140053 | mitochondrial gene expression | 158 | 76 | 0.408 | 1.889 | <0.001 | 0.046 |
| GO:0043583 | ear development | 152 | 43 | -0.384 | -1.756 | 0.002 | 0.057 |
| GO:0050866 | negative regulation of cell activation | 118 | 44 | -0.412 | -1.812 | <0.001 | 0.059 |
| GO:0051339 | regulation of lyase activity | 40 | 12 | -0.510 | -1.844 | <0.001 | 0.059 |
| GO:0060231 | mesenchymal to epithelial transition | 14 | 7 | -0.629 | -1.747 | 0.008 | 0.059 |
| GO:0070371 | ERK1 and ERK2 cascade | 247 | 77 | -0.363 | -1.765 | <0.001 | 0.061 |
| GO:0036314 | response to sterol | 24 | 9 | -0.538 | -1.756 | 0.002 | 0.062 |
| GO:0048144 | fibroblast proliferation | 69 | 29 | -0.429 | -1.731 | <0.001 | 0.063 |
| GO:0070972 | protein localization to endoplasmic reticulum | 135 | 65 | -0.409 | -1.857 | <0.001 | 0.063 |
| GO:0009410 | response to xenobiotic stimulus | 202 | 66 | -0.373 | -1.768 | <0.001 | 0.063 |
| GO:0072089 | stem cell proliferation | 94 | 25 | -0.412 | -1.734 | <0.001 | 0.064 |
| GO:0009451 | RNA modification | 137 | 45 | 0.393 | 1.775 | <0.001 | 0.122 |
| GO:0072512 | trivalent inorganic cation transport | 32 | 10 | 0.498 | 1.697 | 0.008 | 0.196 |
| GO:1902600 | proton transmembrane transport | 121 | 40 | 0.365 | 1.598 | 0.002 | 0.234 |
| GO:0006984 | ER-nucleus signaling pathway | 40 | 17 | 0.445 | 1.609 | 0.008 | 0.245 |
| GO:0006414 | translational elongation | 131 | 39 | 0.344 | 1.549 | 0.004 | 0.258 |
| GO:0010586 | miRNA metabolic process | 23 | 5 | 0.491 | 1.553 | 0.022 | 0.272 |
| GO:0010257 | NADH dehydrogenase complex assembly | 58 | 34 | 0.403 | 1.566 | 0.006 | 0.272 |
| GO:0006399 | tRNA metabolic process | 173 | 56 | 0.347 | 1.612 | <0.001 | 0.279 |

ES = Enrichment score; NES = Normalized enrichment score.

TABLE S2: **Summary of gene set enrichment analysis for the first significant latent variable (PLS2).**

| **Gene Set** | **Description** | **Size** | **Leading Edge Number** | **ES** | **NES** | ***P*-Value** | **FDR** |
|---|---|---|---|---|---|---|---|
| GO:0140053 | mitochondrial gene expression | 158 | 83 | 0.428 | 1.949 | <0.001 | 0.024 |
| GO:0010257 | NADH dehydrogenase complex assembly | 58 | 33 | 0.507 | 1.967 | <0.001 | 0.032 |
| GO:0033108 | mitochondrial respiratory chain complex assembly | 88 | 47 | 0.439 | 1.842 | <0.001 | 0.067 |
| GO:0000959 | mitochondrial RNA metabolic process | 40 | 15 | 0.498 | 1.782 | <0.001 | 0.080 |
| GO:0097503 | sialylation | 18 | 8 | 0.592 | 1.763 | 0.007 | 0.082 |
| GO:1902600 | proton transmembrane transport | 121 | 46 | 0.407 | 1.794 | <0.001 | 0.088 |
| GO:0006414 | translational elongation | 131 | 58 | 0.393 | 1.733 | <0.001 | 0.095 |
| GO:0061614 | pri-miRNA transcription by RNA polymerase II | 36 | 14 | -0.445 | -1.712 | 0.005 | 0.107 |
| GO:0048144 | fibroblast proliferation | 69 | 29 | -0.395 | -1.718 | 0.003 | 0.109 |
| GO:0031128 | developmental induction | 22 | 12 | -0.515 | -1.719 | 0.007 | 0.116 |
| GO:0009636 | response to toxic substance | 396 | 123 | -0.303 | -1.692 | <0.001 | 0.120 |
| GO:0032103 | positive regulation of response to external stimulus | 204 | 73 | -0.323 | -1.724 | <0.001 | 0.124 |
| GO:0007588 | excretion | 48 | 12 | -0.411 | -1.681 | <0.001 | 0.124 |
| GO:0007271 | synaptic transmission, cholinergic | 18 | 6 | -0.494 | -1.557 | 0.026 | 0.135 |
| GO:0070371 | ERK1 and ERK2 cascade | 247 | 78 | -0.324 | -1.724 | <0.001 | 0.136 |
| GO:0003205 | cardiac chamber development | 134 | 44 | -0.318 | -1.558 | <0.001 | 0.138 |
| GO:0045730 | respiratory burst | 20 | 11 | -0.483 | -1.562 | 0.022 | 0.141 |
| GO:0006984 | ER-nucleus signaling pathway | 40 | 15 | 0.455 | 1.640 | 0.009 | 0.213 |
| GO:0010586 | miRNA metabolic process | 23 | 5 | 0.474 | 1.508 | 0.039 | 0.427 |
| GO:0006813 | potassium ion transport | 188 | 78 | 0.316 | 1.472 | 0.001 | 0.440 |

ES = Enrichment score; NES = Normalized enrichment score.
